# Multiplex Solid-Phase RPA Coupled CRISPR-Based Visual Detection of SARS-CoV-2

**DOI:** 10.1101/2021.11.02.21265831

**Authors:** Xiaochen Qin, Yuyuan Zhou, Ratul Paul, Yue Wu, Yaling Liu

## Abstract

COVID-19 has challenged the world’s public health and led to over 4.5 million deaths. A rapid, sensitive, and cost-effective point-of-care virus detection device is crucial to the control and surveillance of the contagious severe acute respiratory syndrome coronavirus-2 (SARS-CoV-2) pandemic. Here we demonstrate a solid phase isothermal recombinase polymerase amplification coupled CRISPR-based (spRPA-CRISPR) assay for on-chip multiplexed, sensitive and visual COVID-19 DNA detection. By targeting the SARS-CoV-2 structure protein encoded genomes, two specific genes were simultaneously detected with the control sample without cross-interaction with other sequences. The endpoint signal can be directly visualized for rapid detection of COVID-19. The amplified target sequences were immobilized on the one-pot device surface and detected using the mixed Cas12a-crRNA collateral cleavage of reporter released fluorescent signal when specific genes were recognized. The system was tested with samples of a broad range of concentrations (20 to 2×10^5^ copies) and showed analytical sensitivity down to 20 copies per reaction. Furthermore, a low-cost LED UV flashlight (∼$12) was used to provide a visible SARS-CoV-2 detection signal of the spRPA-CRISPR assay which could be purchased online easily. Thus, our platform provides a sensitive and easy-to-read multiplexed gene detection method with the capacity to specifically identify low concentration genes. Similar CRISPR biosensor chips can support a broad range of applications such as HPV DNA detection, influenza SARS-CoV-2 multiplex detection, and other infectious disease testing assays.

**Highlights:**

- A commercially available UV flashlight excited CRISPR-Cas12a based visual sensor combined with solid-phase RPA
- The developed assay could detect SARS-CoV-2 down to 20 copies with no signal crosswalk
- This multiplexed on-chip virus detection system would provide great insight for detecting and differentiating SARS-CoV-2 from other viral infections with a single sample simultaneously.

## 1. Introduction

The outbreak of the SARS-CoV-2, also known as COVID-19, has rapidly spread to produce a global pandemic and caused a huge number of deaths worldwide. The lack of rapid accessible and accurate molecular diagnostic testing methods has hindered the COVID-19 testing response as the pandemic quickly spread. Currently, the global standard approach approved for COVID-19 diagnosis is the quantitative reverse transcription polymerase chain reaction (RT-qPCR)[1], [2]. However, because of the rapid growth of the infected population, there is a global shortage of RT-qPCR assay kits as well as their associated reagents [3]. The operation of RT-qPCR also requires highly skilled lab personnel and specialized equipment. The detection of the virus using RT-qPCR usually takes 4-6 hours, and the shipping of samples to the local laboratories has delayed the turnaround diagnosing time to >24 hours [4]. Therefore, the availability of rapid, high sensitivity, multiplexed [5], [6] and precision point-of-care tools [7], [8] for diagnostic technologies while reducing the need for expertise and infrastructure are desired.

New isothermal nucleic acid amplification approaches including recombinase polymerase amplification (RPA) and loop-mediated isothermal amplification (LAMP) may meet the current demand of reducing the need for lab equipment. The LAMP technology typically requires six primers with the reaction temperature at 50–72 °C, while RPA appears to have some attractive features as it requires only two primers with a faster reaction time and optimally works under lower working temperatures ranging from 37°C to 42°C [9], [10]. However, in the RPA reaction, the formation of primer-dimers could cause the production of the non-specifically amplified by-products with the presence of low concentration target DNA [11]. Solid-phase amplification has inherent advantages of RPA and has been used to address the drawbacks of RPA. Solid-state RPA is a detection technique developed based on RPA where the primers are pre-immobilized to the substrate [12]. During the amplification, one of the primers is covalently immobilized on a specific surface, the target amplified with elongation of the primers in both liquid phase and solid phase simultaneously. Therefore, the formation of primer-dimers can be successfully minimized, thus reducing the formation of randomly sequenced by-products [13]. Previously reported multiplex testing platform [14] has pre-immobilized the LAMP primers by drying down on the microfluidic channel surface, which enabled a one-step amplification reaction by reconstituting the primers into the solution. Solid-phase RPA could easily be adaptable to multiplexed capacity by immobilizing different surface primers in an array format on-chip [12] and also allow further detection steps on the immobilized amplified DNA targets.

RT-RPA or RT-LAMP have been used to detect COVID-19 [9], [14]–[16]. The detection sensitivity of these isothermal DNA amplification assays could be further enhanced by coupling with clustered regularly interspaced short palindromic repeats (CRISPR) system [4], [17], [18]. Cas proteins can be activated upon binding with target double-stranded DNA or RNA and unleash nonspecific single-stranded DNA or RNA cleavage activity by AsCas12a (Cpf1) proteins [13], [19]–[22]. When combined with DNA amplification, the CRISPR-based assay typically visualized the results on the lateral flow strips or fluorescent readers with good sensitivity [4], [23]. However, these CRISPR-based DNA detection methods typically require multiple manual operations which potentially increase the risk of contaminations while transferring products between wells.

Herein, we report the development of the solid-phase RPA coupled with a CRISPR-based (spRPA-CRISPR) multiplex detection system to sensitively detect the SARS-CoV-2 related genes. The detection result at the end of reactions could be directly visualized under a commercially available UV flashlight. The detection mechanism is illustrated in Fig. 1. To create this system, a single cartridge mixture of CRISPR RNA (crRNA) probes for E and N genes were applied to specifically detect the thiol-group immobilized and amplified gene targets within one-well without signal crosswalk. Further, we quantitatively evaluated the range and detection limit of spRPA-CRISPR assay targeting COVID-19 genes. Last, the endpoint on-chip assay reads were stimulated by the low-cost UV flashlight and visualized by the naked eye. This assay platform exhibited a lower limit of detection (LOD) of 20 copies with a broad testing range at a constant temperature of 37 °C (compared with the traditional RPA detection methods. The use of spRPA can largely simplify the chip design and the handling procedures by performing all the reactions within one-well which may easily be adaptable for sensitive POC multiplex virus detection.

**Figure 1.**
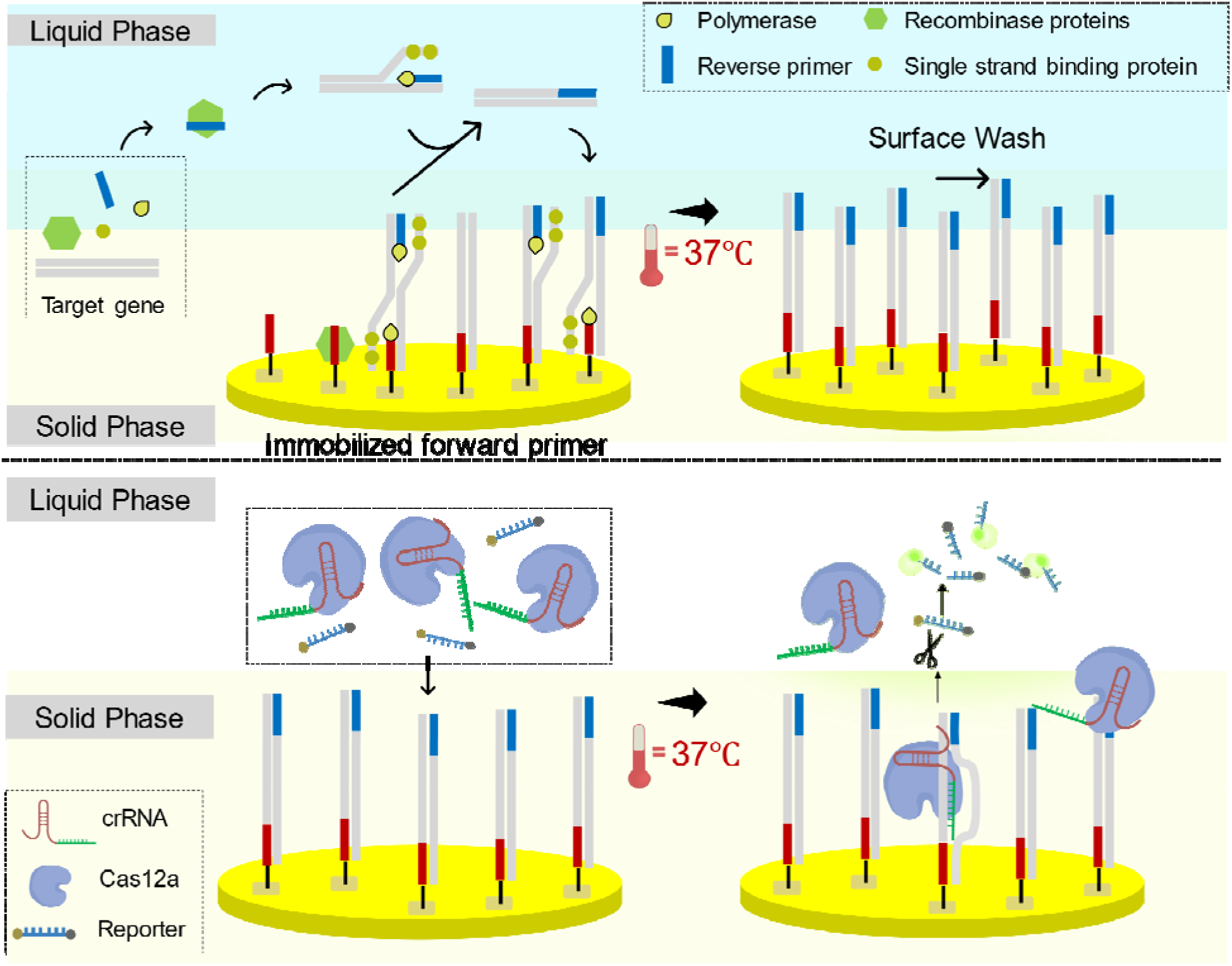
The schematic of solid-phase RPA (top) and CRISPR-based SARS-CoV-2 colorimetric detection (bottom)

## 2. Materials and Methods

### 2.1. Materials and primer design

The TwistAmp™ basic kit was purchased from TwistDx (Babraham, UK, www.twistdx.co.uk) for RPA reactions. Primers and crRNA were designed by following the TwistDx kit instruction and IDT protocol. Oligonucleotides (primers and crRNAs) and ssDNA-FQ reporters were purchased from IDT (Integrated DNA Technologies, US). The DNA template using COVID-19 N gene plasmid (2019-nCoV_N_Positive Control, IDT) and E gene (2019-nCoV_E_Positive Control, IDT). The RPA amplified DNA products were purified using QIAquick PCR Purification Kit (Qiagen) and were confirmed by agarose gel electrophoresis. The Sylgard 184 PDMS elastomer was mixed with the curing agent at a ratio of 10 and poured into a plastic petri dish at 30 mm height. After curing, the PDMS was cut into squares (1.0 × 1.0 × 0.3 cm) with a 0.5 cm diameter hole punched in the middle.

**Table 1.**
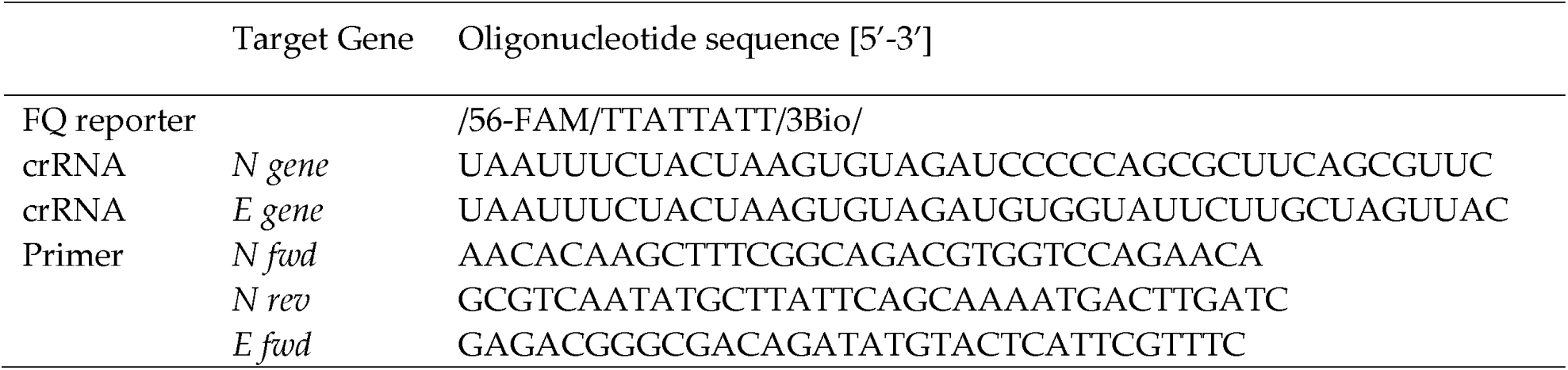
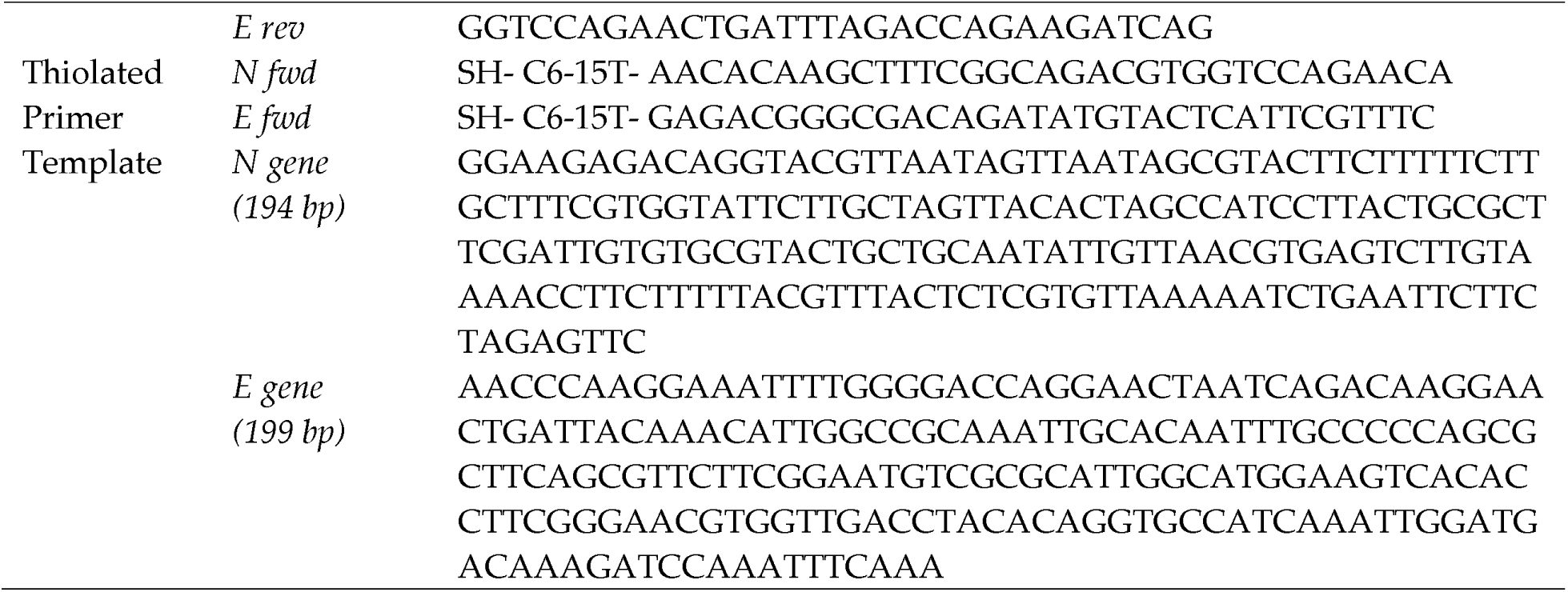
Probes and target sequences

### 2.2. Primer design and probe verification by RPA assay

The materials for RPA reaction including buffers and reagents were supplied in TwistDx kit. The primers were first evaluated with RPA reaction for further solid-phase reaction. The RPA assay was tested in a 50 μL reaction volume containing 480 nM of forward and reverse primers, 1x rehydration buffer, 2000 copies of genomic DNA strand. This reaction mixure was added to one tube of freeze-dried TwistAmp Basic reaction pellet and 14 mM Magnesium acetate was then added to the mix to start the reaction. The samples were incubated at 37 °C (for 20 min. Amplicons generated were then cleaned of DNA-binding proteins using PCR purification columns to be visualized using 2% agarose gel electrophoresis. The amplified products were detected using ethidium bromide (Thermo Fisher).

### 2.3. CRISPR Cas12 cleavage reaction and LED readout

For the CRISPR assay, the crRNA of E gene or N gene were first assembled into Cas12a-crRNA complex with a total of 200 nM of As Cas12a (Cpf1) Nuclease (Cat. 1081068, IDT) was preincubated with 250 nM g-RNA in 1x binding buffer (20 mM Tris-HCl, 100 mM KCl, 5 mM MgCl2, 1 mM DTT, 5% glycerol) at 37 °C for 30 min. The reactions were performed separately for the N gene and E gene. After forming the Cas12a-crRNA complex, the FQ reporter was added to the reaction product at a final concentration of 500 nM. The complexes were placed on ice for immediate use or stored at 4 °C for up to 24 hours. After the RPA amplification step, 2 μL of amplicons and 20 μL of CRISPR reaction complexes were mixed with 78 μL of 1x binding buffer to allow the Cas12a trans-cleavage assay to proceed. After 10 min incubation at 37 °C, the samples were ready to read by plate reader using blue excitation light or visualized under ultraviolet (UV) LED flashlight (Vansky).

### 2.4. Solid-Phase RPA reaction

#### Gold pattern preparation and device assembly

The gold platform was designed with a set of three identical circular patterns for thiol-gold immobilization. Each pattern was designed with 300 μm in diameter, the gap between the electrodes is 250 μm and the outer electrode is 25 μm wide and is open on one side [24]. The metal deposition of the designed pattern was fabricated using standard lithography protocol on a 76.2mm soda-lime glass wafer with 500 ⍰ titanium layer and 2000 ⍰ gold layer. The metallization patterns were defined using the lift-off technique to remove the extra photoresist. After fabricating the gold pattern, three PDMS wells were autoclaved and placed on top of each pattern parallelly for primer immobilization.

#### Immobilization of forward primer

The forward primer was designed with a poly-15T at the 5’ end of the probe as a vertical spacer. 6-mercapto-1-hexanol (MCH, Sigma-Aldrich) is a commonly used, stable, and cost-effective lateral spacer. The MCH was optimized to be mixed with forward primers with a 1:10 ratio for surface immobilization [12]. Reduction for oligos with thiol modification has followed the protocol provided by IDT [25]. 1 μL of 10 μM thiolated forward primer for E gene or N gene with 100 μM MCH were prepared in 1M KH2PO4 (Sigma-Aldrich), the mixtures were added into separate PDMS chambers for immobilization on gold surface. The coating was carried out in a humid chamber at room temperature for 20 hr. The gold surface was then washed to remove non-specific immobilization of DNA by (1) rinsed in Milli-Q water for 5 min. (2) dried in the stream of N2. (3) washed for 5 min in PBS, Tween 20, and Milli-Q. Afterward, the surface was blocked and prevented non-specific binding from subsequent steps with Denhardt’s solution (Thermo Fisher) at room temperature for 20 min. As a final step, the chamber was washed with PBS and Milli-Q water for 5 min and dried with the nitrogen stream. The forward primer immobilized surface could be left under vacuum [12]or kept in 4 ([11] until needed.

#### 2.4.3. Solid-Phase RPA reaction on gold surface

Solid-phase RPA was performed by mixing 2.4 μL of 10 μM reverse primer, 13.2 μL of 10 nM DNA template, 1x rehydration buffer, DNase free water mixed with lyophilized pellet from TwistAmp Basic kit. The master mix of all reagents except for magnesium acetate was prepared and pipetted to transfer into PDMS wells. Finally, add/mix 2.5 μL of 280 mM magnesium acetate gently into each PDMS well as the last step to initiate the amplification. The samples were left to incubate at 37 °C for 60 min.

#### 2.4.4. One-well spRPA-CRISPR detection with LED readout

The CRISPR reagent preparation was described in the “CRISPR reaction and LED readout” section. The spRPA amplified surface was rinsed twice with PBS. The crRNA of N gene or E gene were first prepared with CRISPR-Cas12a protein and formed crRNA-CRISPR complex, then mixed with fluorophore-quencher (FQ) reporter, and 1x binding buffer to bring the final reaction volume to 100 μL. After incubating for 10 min at 37 °C, the samples were ready to detect under an LED flashlight or plate reader. The visible colorimetric detection result was analyzed using ImageJ software by splitting the green channel and measuring the fluorescent intensity [26].

## 3. Results and discussions

### 3.1 Evaluation of liquid phase RPA coupled CRISPR-Cas12-based assay for COVID-19 diagnosis

To assess the capability of amplifying the target genes of the SARS-CoV-2 in the solid-phase RPA, the designed primers were first verified using liquid phase RPA reaction. Liquid RPA was conducted using the standard technique provided by TwistDx. The liquid RPA performance was proved effective with the primers and genomic DNA template binding and amplified target sequences. The successful amplification of the target N gene and E gene were verified by running agarose gel, as shown in Fig.2 (a).

Next, we evaluated the performance of the CRISPR-Cas12a assay on the RPA products. The assay is considered positive if the fluorescent signal is detected for both E and N genes. The crRNAs were specifically designed for E and N genes separately. The CRISPR-Cas12a enzyme complexed crRNA will specifically detect the genes that carry complementary sequences from the RPA product. The fluorescent signal is generated when the Cas12a-crRNA complex recognizes the predefined target sequences and collaterally cleaves the surrounding FQ reporter molecules. The flashlight allows sensitive and quantitative readout of the CRISPR-Cas12-based assay. In Fig. 2 (b), the fluorescent signal for both E and N genes was detected under the excitation of a UV flashlight or fluorescent plate scanner. The visible signal was directly observed under the flashlight and imaged using a regular cellphone camera. The image was split into the red, blue and green color channels using ImageJ. The fluorescent signal was analyzed from both the green color channel of the visual detection result and the measurement from the plate scanner. As shown in Fig.2 (b), fluorescent intensity measurements for both UV flashlight visual detection and plate reader agreed with each other.

**Figure 2.**
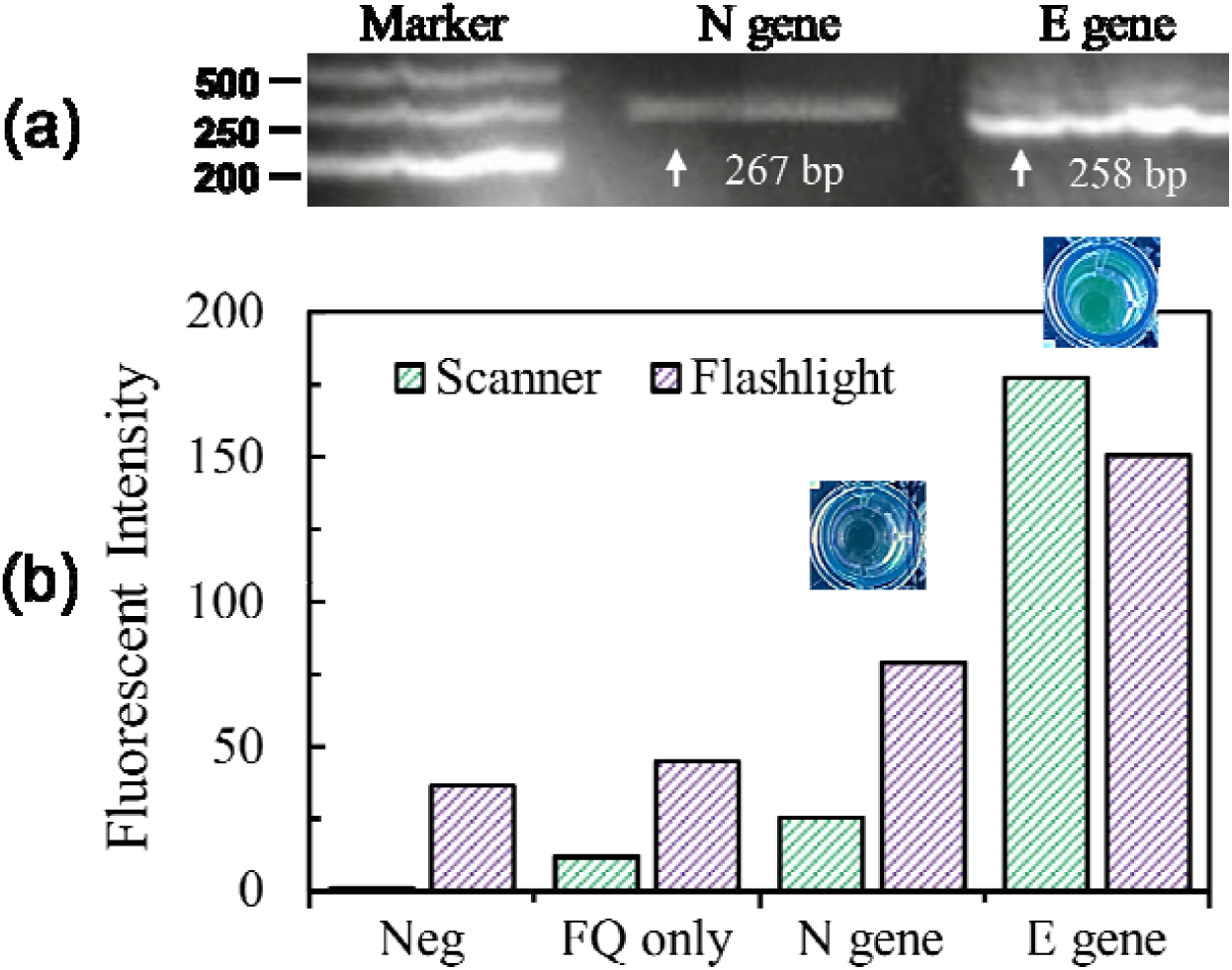
Primers and probe verification with RPA-based CRISPR detection assay (a) Agarose gel (2%) illustration of liquid-phase RPA product of N gene and E gene. (b) Fluorescence measurement using plate reader and UV flashlight.

### 3.2 Evaluation of solid-phase RPA-CRISPR assay for COVID-19 diagnosis

As a proof-of-concept for the multiplexed isothermal spRPA-CRISPR mechanism, the format of the DNA amplification method consisted of the patterned gold surface as solid-phase, the bulk solution as liquid-phase, and the interface where the thiolated forward primer was bond covalently with the gold layer supported by a glass substrate. The immobilized primers facilitate the elongation by the polymerase reaction by vertical poly-15T spacer and lateral MCH spacer, while the vertical spacer projects the primer from the surface and the lateral spacer avoids undesirable interactions between neighboring primers [12]. In Fig. 1, the process of spRPA has been described on both solid and liquid phases: on the solid phase, the immobilized forward primers serve as a starting point and elongate the strand following the polymerase. In the liquid phase, the DNA first elongates along the reverse primer and then releases to hybridize specifically to the immobilized forward primers[13]. To perform the CRISPR assay with the DNA amplified strains only elongated from the immobilized primers, the surface was rinsed twice with PBS before introducing the Cas12a-crRNA complex and FQ reporter to the system.

The system was then further transferred to a solid-phase multiplexing platform. The gene-specific amplification along the surface-immobilized primers was achieved within 60 min. The CRISPR-Cas-based assay was performed as an extended step to enhance the detection target viral genome amplicon signal and showed a visible signal within 10 min incubation. To identify the linear range and LOD of the optimized assay, the target plasmid templates were tested in the concentration range of 0 copies to 20000 copies per μL. The results were shown in Fig. 3. (a), distinct fluorescent signals were shown between the negative control group where primers were not amplified with un-cleaved FQ reporter and the sample groups for the LOD test. A strong fluorescent signal was detected for both UV flashlight excited visualized signal and fluorescent imaging scanner.

**Figure 3.**
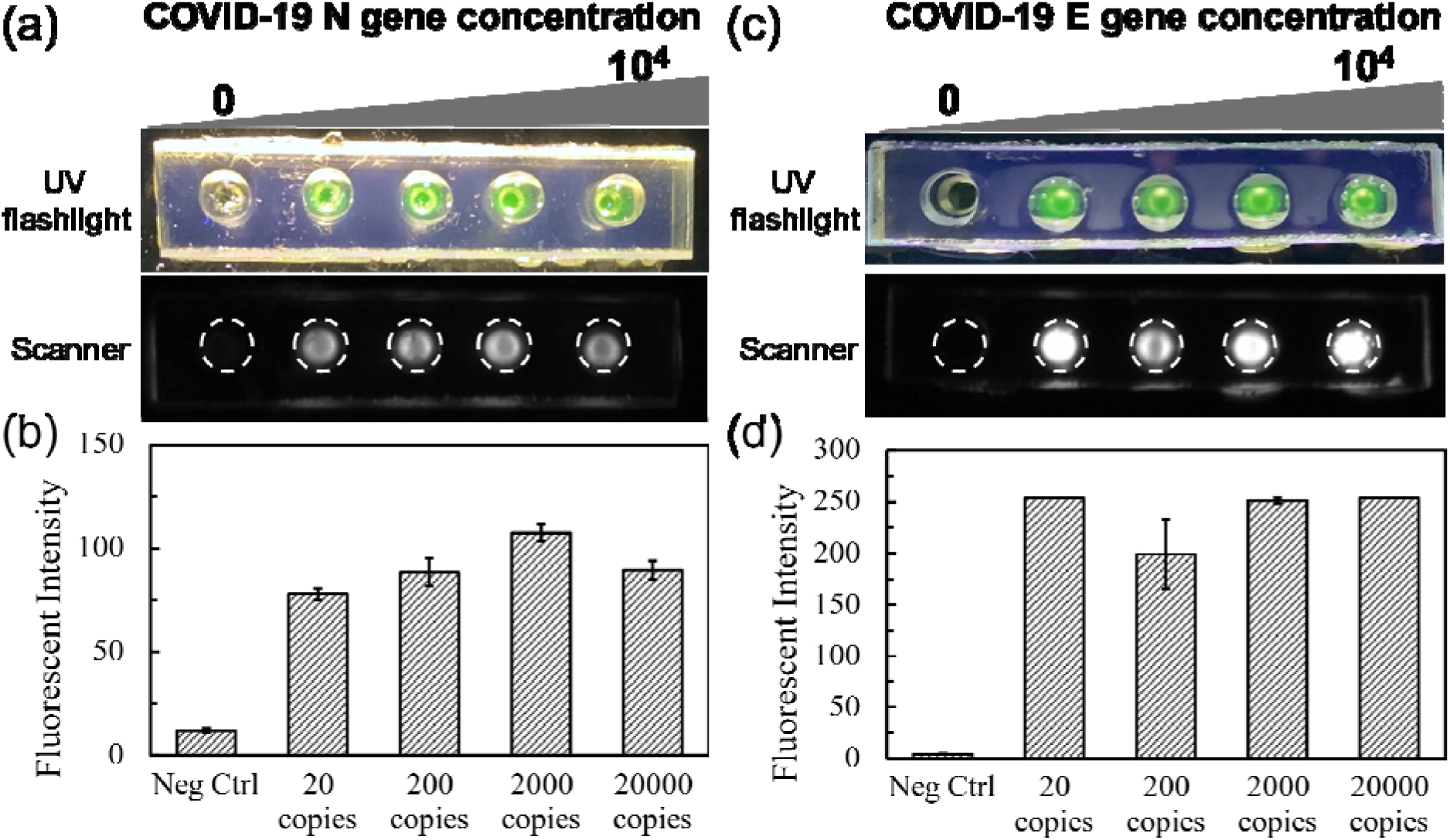
Sensitivity of spRPA-based CRISPR assay. (a) Fluorescence signal excited with LED flashlight and fluorescent imaging system of N gene (0, 20, 200, 2000, 20000 copies). (b) Fluorescence signal measurement of N gene. (c) Fluorescence signal excited with UV flashlight and fluorescent imaging system of E gene (0, 20, 200, 2000, 20000 copies). (d) Fluorescence signal measurement of E gene.

Next, we evaluated the specificity of our sp-RPA-based CRISPR system for the multiplexed detection ability to simultaneously detect the N gene and E gene. We added Cas12a-crRNA of E gene or N gene separately into E gene or N gene amplified well to test if the CRISPR assay could specifically detect the amplified DNA product in Fig.4. When the Cas12a-crRNA complex finds their complementary genes, the wells display strong fluorescent signals which indicate successful detection, and while the Cas12a-crRNA complex was not able to find their target sequences, the wells remain dark. The negative wells exhibit a low level of background fluorescent signal presumably from the weak autofluorescence from FQ reporter and low level of non-specific primer-dimer existed in the DNA amplification. As shown in Fig. 4 (a), the Cas12a-crRNA complexes of the E gene were added separately into the spRPA product amplified from the E gene primer and N gene primers, which separately resembled the left well of both Fig.4 (b) and (c). In Fig.4 (d), both N gene and E gene crRNA complexed Cas12a-crRNA were mixed and added separately into the solid phase N gene or E gene amplification well. As shown in Fig. 4, COVID-19 E gene and N gene were identified and distinguished from each other with the gene-specific detection of both targets could be directly visualized using the spRPA-CRISPR assay.

**Figure 4.**
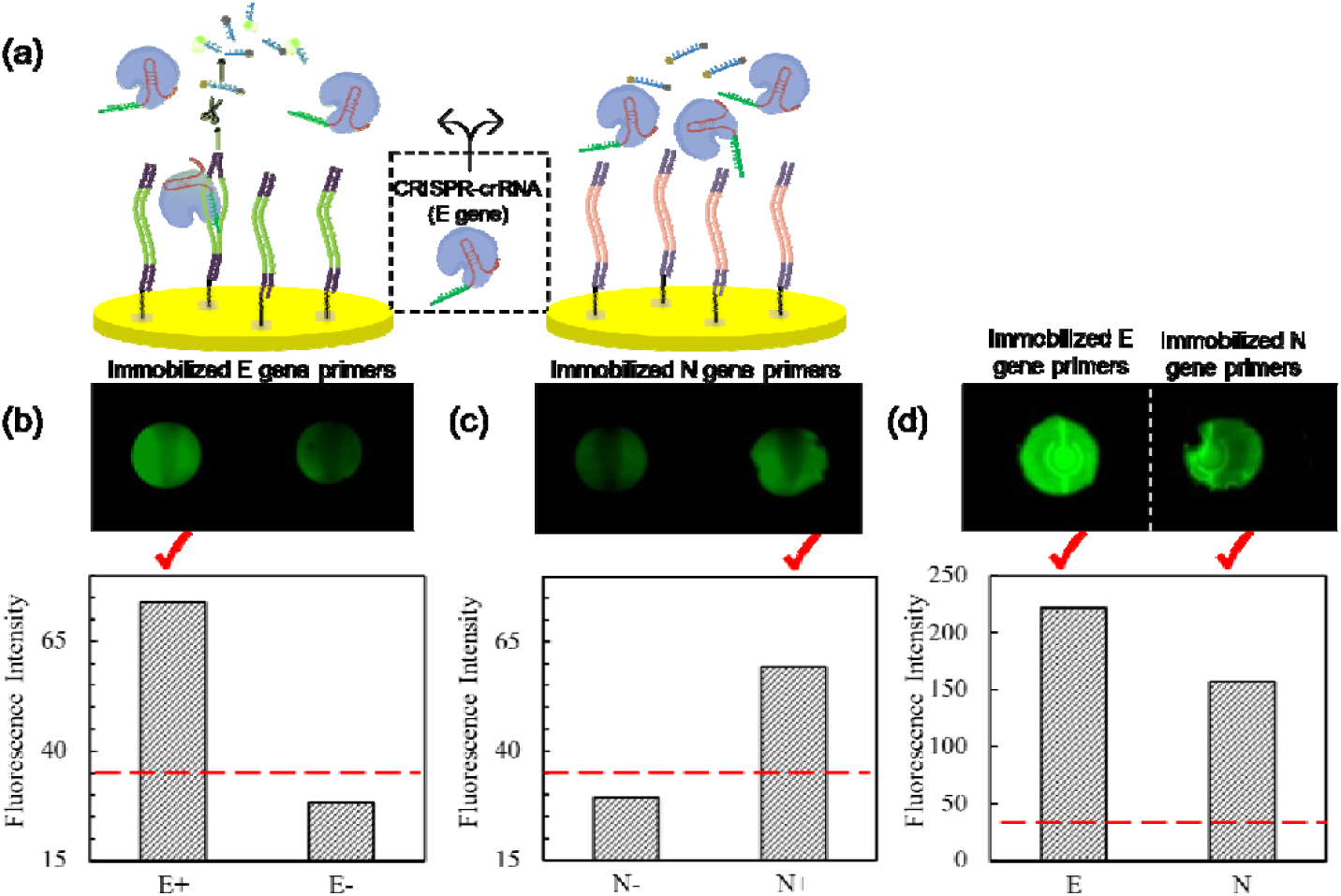
Multiplex detection of COVID-19 genes. (a) Illustration of the gene specificity of CRISPR detection on the spRPA product, the left figure demonstrated the left well in (b) where the Cas12a-crRNA of E gene bind with amplified E gene thus triggered the trans-cleavage of FQ reporter, while the right figure demonstrated the left well in (c) where the Cas12a-crRNA of E gene were not activated. (b) CRISPR detection fluorescent image and the fluorescent intensity scan for E gene specificity test: spRPA amplified E gene on the well bottom; Left well: detected using Cas12a-crRNA of E gene; Right well: detected using Cas12a-crRNA of N gene. (c) The CRISPR detection fluorescent image and the fluorescent intensity scan for N gene specificity test: spRPA amplified N gene in both wells. Left well: detected using Cas12a-crRNA of E gene; Right well: detected using Cas12a-crRNA of N gene. (d) The single cartridge mixed Cas12a-crRNA of both E gene and N gene detected immobilized E gene (left) and immobilized N gene (right)

To further demonstrate the PoC diagnostic application, we analyzed the visual detection signal under UV flashlight excitation. In the on-chip spRPA-CRISPR assay, the N gene, E gene primers and a non-template negative control were prepared separately within each well. The CRISPR assay was performed using a single cartridge that mixed crRNA of both N gene and E gene on the spRPA product that amplified from 20 copies/μL for each of the target genes. The fluorescent signals were detectable after 10 mins of incubation and visualized under the UV flashlight. The process of the spRNA-CRISPR assay workflow including signal visualization and fluorescent signal analysis was shown in Fig. 5.

**Figure 5.**
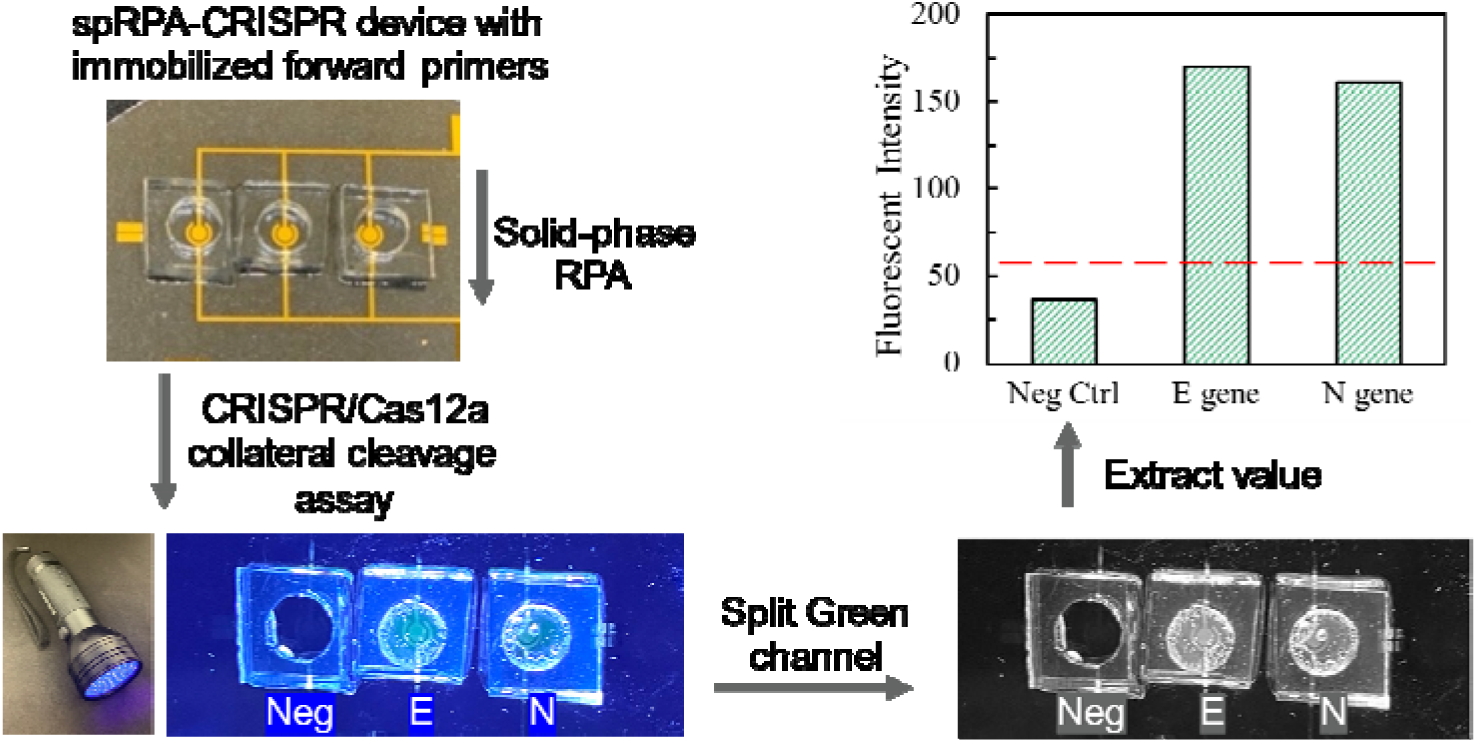
Workflow of the spRPA-CRISPR chip for DNA detection. Fabricated prototype device bound with PDMS chamber and use LED flashlight to observe the endpoint fluorescent signal. Image were taken using a smartphone camera and analyzed the intensity by splitting green channel using ImageJ. Single cartridge reaction containing Cas12a-crRNA complex with both E gene and N gene test on the spRPA amplified target DNA templates with negative control with no amplified gene, E gene, and N gene (left to right).

## 4. Discussions

In this study, we have demonstrated a sensitive, highly specific, and cost-effective multiplexed spRPA-CRISPR assay for the detection of SARS-CoV-2, where the end-point results were directly visualized. This method is, to the best of our knowledge, the first system that allows CRISPR-based assay to specifically detect the “pre-programmed” N gene, E gene, and control panels by covalently immobilizing and amplifying the target DNA sequence on the chip surface. Our system can detect single or multiple specific nucleic acid targets simultaneously with flexible solid-phase multiplexed CRISPR detection, which is valuable for multiplex detection of infectious diseases. The system could successfully detect both E and N genes of 20 template copies per reaction. More importantly, the CRISPR reaction product can be directly visualized under the excitation of a low-cost commercially available LED flashlight which eliminates the need for extra lateral flow assay step and simplifies the detection process [4].

Future improvement and development are to integrate our spRPA-CRISPR system into a disposable multiplexed microfluidic system, enabling an integrated sample-to-result detection system. The microfluidic channel with pre-immobilized primers could be dried with a nitrogen beam and stored in vacuum packing until use, while other reagents for our spRPA-CRISPR detection system could be stored in lyophilized form at room temperature. Therefore, the system will require only a small space and eliminate the need for cold chains for shipping and storage, while also enabling a point-of-care setting. With the endpoint signal directly visualized by naked eyes, it is also possible to record the endpoint signal directly using the smartphone camera by taking the advantage of using the 3D printed holder assembled with smartphone and optical filters. The smartphone cameras can be further programmed to take fluorescent images can be quickly transmitted to a cloud server for results analysis. By using the server to perform data normalization and analysis, quantitative test results will be made available to clinical doctors and public health organizations in a timely manner. We believe that this spRPA-CRISPR system offers the potential to rapidly screen COVID-19 or other viral transfection diseases with high sensitivity and specificity.

## Data Availability

All data produced in the present work are contained in the manuscript

## 5. Acknowledgement

This work was supported by National Science Foundation grant CBET 2039310, and National Institute of Health grant R01HL131750.

## 6. Author Contributions

Xiaochen Qin designed and constructed the solid-phase RPA based CRISPR detection system. Yuyuan Zhou designed primers and CRISPR probes, Ratul Paul performed the lithography, Yue Wu designed the surface gold patterns. Xiaochen Qin wrote the manuscript. Yuyuan Zhou, Ratul Paul, Yue Wu and Yaling Liu revised the manuscript. Yaling Liu developed the idea.

